# Comparison of synthesized and acquired high *b*-value diffusion-weighted MRI for detection of prostate cancer

**DOI:** 10.1101/2023.02.17.23286100

**Authors:** Karoline Kallis, Christopher C. Conlin, Allison Y. Zhong, Troy S. Hussain, Aritrick Chatterjee, Gregory S. Karczmar, Rebecca Rakow-Penner, Anders Dale, Tyler Seibert

## Abstract

**Background:** High *b*-value diffusion-weighted images (DWI) are used for detection of clinically significant prostate cancer (csPCa). To decrease scan time and improve signal-to-noise ratio, high *b*-value (>1000 s/mm^2^) images are often synthesized instead of acquired.

**Purpose:** Qualitatively and quantitatively compare synthesized DWI (sDWI) to acquired (aDWI) for detection of csPCa.

**Study Type:** Retrospective

**Subjects:** 151 consecutive patients who underwent prostate MRI and biopsy.

**Sequence:** Axial DWI with *b*=0, 500, 1000, and 2000 s/mm^2^ using a 3T clinical scanner using a 32-channel phased-array body coil

**Assessment:** We synthesized DWI for *b*=2000 s/mm^2^ via extrapolation based on monoexponential decay, using *b*=0 and *b*=500 s/mm^2^ (sDWI_500_) and *b*=0, *b*=500, and *b*=1000 s/mm^2^ (sDWI_1000_). Differences between sDWI and aDWI were evaluated within regions of interest (ROIs). The maximum DWI value within each ROI was evaluated for prediction of csPCa. Classification accuracy was also compared to Restriction Spectrum Imaging restriction score (RSIrs), a previously validated biomarker based on multi-exponential DWI.

**Statistical Tests:** Discrimination of csPCa was evaluated via area under the receiver operating characteristic curve (AUC). Statistical significance was assessed using bootstrap difference (two-sided α=0.05).

**Results:** Within the prostate, mean ± standard deviation of percent mean differences between sDWI and aDWI signal were -46±35% for sDWI_1000_ and -67±24% for sDWI_500_. AUC for aDWI, sDWI_500,_ sDWI_1000_, and RSIrs within the prostate 0.62[95% confidence interval: 0.53, 0.71], 0.63[0.54, 0.72], 0.65[0.56, 0.73] and 0.78[0.71, 0.86], respectively. When considering the whole field of view, classification accuracy and qualitative image quality decreased notably for sDWI compared to aDWI and RSIrs.

**Data Conclusion:** sDWI is qualitatively comparable to aDWI within the prostate. However, hyperintense artifacts are introduced with sDWI in the surrounding pelvic tissue that interfere with quantitative cancer detection and might mask metastases. In the prostate, RSIrs yields superior quantitative csPCa detection than sDWI or aDWI.

## Introduction

Diffusion-weighted imaging (DWI) is a critical component of multiparametric MRI for the detection and characterization of clinically significant prostate cancer (csPCa)^1^. The degree of diffusion-weighting in DWI is indicated by the *b*-value, with higher *b*-values corresponding to images with less signal where water in tissues diffuses more rapidly^2^. High *b*-values are used for their greater tumor conspicuity and detection of even small lesions^3^. The Prostate Imaging – Reporting and Data System (PI-RADS v2.1) recommends the acquisition of high *b*-values (1400-2000 s/mm^2^) for lesion detection, without precisely defining an optimal value for csPCa^4^. While clinically valuable, high *b*-values require more scan time and suffer from low signal-to-noise ratio (SNR) and increased susceptibility to artifacts due to microscopic motion or small fluctuations in local magnetic field. One common solution, permitted by PI-RADS, is to synthesize high *b*-value images by extrapolating signal from acquired low *b*-value images using a mono-exponential model^1,5^. However, mono-exponential models do not adequately represent restricted diffusion in complex tissues^6,7^, possibly calling into question the accuracy of synthesized images.

More advanced DWI models have been developed to better account for tissue microstructure, including intravoxel incoherent motion imaging^8,9^, diffusion kurtosis imaging^10,11^, Vascular, Extracellular, and Restricted Diffusion for Cytometry in Tumor (VERDICT)^12–14^, hybrid multidimensional MRI (HM-MRI)^15–18^, and Restriction Spectrum imaging (RSI)^12,19^. In RSI, the diffusion signal is modeled as a weighted sum of different compartments representing different tissue types^19,20^. The RSI restriction score (RSIrs) is based on the model coefficient for the most restricted diffusion compartment and has been shown to be a useful biomarker for the detection of csPCa^20–22^.

Studies have yielded contradicting results on whether synthesized *b*-values are clinically interchangeable with acquired DWI (aDWI) images. Liu *et al*.^23^ compared various models, including the standard mono-exponential, for the detection of csPCa and concluded that non-linear fitting with various *b*-values is superior to simpler models. In contrast, other studies reported better image quality for synthetic DWI (sDWI) with a similar tumor detection rate in comparison to acquired DWI^5,24–27^.

In this study, we qualitatively and quantitatively analyzed the differences between acquired and synthesized high *b*-value images for detection of csPCa. Further, we compared the detection rate of csPCa between DWI and RSIrs.

## Materials and Methods

### Patient Cohort

This retrospective study was approved by the institutional review board (IRB 805394). 151 men were included in the study who underwent MRI examination between November 2017 and December 2020. These were consecutive patients scanned with the same protocol and for whom results of a prostate biopsy (performed within 180 days of the MRI) were available. Patient characteristics are summarized in **Table 2**.

For all patients, suspicious lesions were contoured per PI-RADS v2.1 by board-certified radiologists using MIM software (MIM Software, Inc; Cleveland, OH). Whole-gland prostate segmentation was performed using OnQ Prostate software (Cortechs Labs, San Diego, CA, USA). Clinically significant prostate cancer (csPCa) was defined as grade group ≥ 2. On biopsy, 86 of the 151 patients were found to have csPCa, while 65 had only benign tissue or grade group 1 cancer. In patients who underwent prostatectomy, grade group was determined per final pathology report.

### MRI Acquisition

All MRI acquisitions were performed on a 3T clinical GE scanner (Discovery MR750, GE Healthcare, Waukesha, WI, USA) using a 32-channel phased-array body coil surrounding the pelvis. Acquisition parameters are summarized in **Table 1**. A single axial DWI volume was acquired for each patient. *T*_*2*_-weighted reference images were acquired for all patients with field of view (FOV) identical to the DWI volume. RSI calculations were performed as described in prior studies ^20–22^.

**Table 1.**
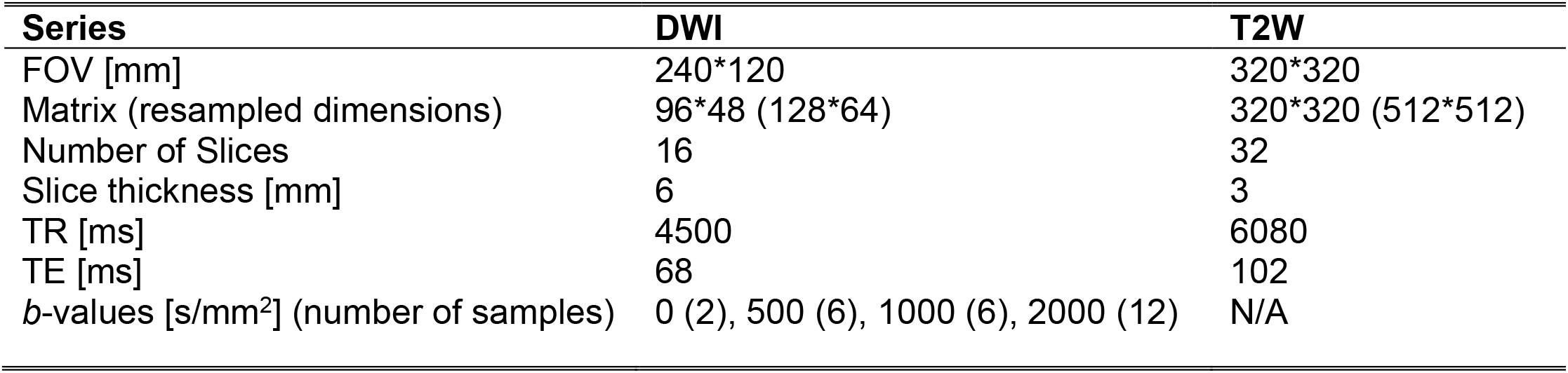
Acquisition parameters for clinical multi-parametric MRI; DWI = diffusion-weighted imaging; T2W = T_2_ weighted MRI

| Series | DWI | T2W |
| --- | --- | --- |
| FOV [mm] | 240*120 | 320*320 |
| Matrix (resampled dimensions) | 96*48 (128*64) | 320*320 (512*512) |
| Number of Slices | 16 | 32 |
| Slice thickness [mm] | 6 | 3 |
| TR [ms] | 4500 | 6080 |
| TE [ms] | 68 | 102 |
| *b*-values [s/mm<sup>2</sup>] (number of samples) | 0 (2), 500 (6), 1000 (6), 2000 (12) | N/A |

**Table 2.** Patient Characteristics range Q_1_-Q_3_= Range between lower first quartile to upper third quartile; csPCa = clinically significant prostate cancer. MRI = magnet resonance imaging; PSA = prostate-specific antigen

| Parameter | Specification | Value |
| --- | --- | --- |
| Number of patients | Total | 151 |
| Age [a] | Median (range Q<sub>1</sub>-Q<sub>3</sub>) | 66 (59-72) |
| Time from MRI to biopsy [d] | Median (range Q<sub>1</sub>-Q<sub>3</sub>) | 16 (1-35) |
| PSA at time of MRI [ng/ml] | Median (range Q<sub>1</sub>-Q<sub>3</sub>) | 7.3 (5.3-10.4) |
| Prostate volume [ml] | Median (range Q<sub>1</sub>-Q<sub>3</sub>) | 45 (34-61) |
| PSA density [ng/ml<sup>2</sup>] | Median (range Q<sub>1</sub>-Q<sub>3</sub>) | 0.16 (0.11-0.25) |
| Best available pathology | Systematic | 7 |
|  | Targeted | 17 |
|  | Systematic and Targeted | 85 |
|  | Prostatectomy | 42 |
| Targeted Biopsy | TRUS Guided | 139 |
|  | Transperineal | 9 |
|  | NaN | 3 |
| PI-RADS Score (csPCa) | I | 0 |
|  | II | 5 (3) |
|  | III | 27 (4) |
|  | IV | 55 (25) |
|  | V | 64 (54) |
| Gleason Grade Group | Benign | 25 |
|  | 1 | 40 |
|  | 2 | 38 |
|  | 3 | 20 |
|  | 4 | 16 |
|  | 5 | 12 |
| Clinical Tumor stage | Negative Biopsy | 25 |
|  | T1c | 94 |
|  | T2a | 13 |
|  | T2b | 11 |
|  | T2c | 8 |

Post-processing of the image data was performed using in-house software in MATLAB (version R2017a, MathWorks, Natick, MA, USA). DWI images were corrected for *B*_*0*_ inhomogeneity distortions, gradient nonlinearity, and eddy currents ^28–30^. Multiple acquired DWI samples at specific *b*-values were averaged together and normalized by median signal intensity of urine in the bladder at *b* = 0 s/mm^2^.

### Synthetic b-value Computation

Synthetic high *b*-value DWI (sDWI) was calculated using the conventional, mono-exponential formula (see below) and using *b*-values up to 500 s/mm^2^ (sDWI_500_) or *b*-values up to 1000 s/mm^2^ (sDWI_1000_).

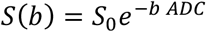

*S(b)* is DWI signal for a given *b*-value, *b. S*_*0*_ is the signal with no diffusion weighting. *ADC* is the apparent diffusion coefficient. sDWI was calculated for *b*=2000 s/mm^2^ to match the acquired high *b*-value DWI (aDWI). To explore the application of sDWI and aDWI for detection of significant cancer lesions outside of the prostate, sDWI and RSIrs were additionally calculated for one representative patient with csPCa and bone metastasis.

### Data Analysis

All data analysis was performed using in-house MATLAB scripts (version R2021a, MathWorks, Natick, MA, USA). Quantitative differences between sDWI and aDWI were estimated by a voxel-wise comparison of the images. Relative deviations were calculated for three different regions of interest (ROIs): prostate, prostate plus a margin of 5 mm, and the whole field of view (FOV) using the following formula:

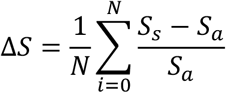

where S_s_ is the synthetic signal intensity, S_a_ the acquired signal and N the number of voxels in the considered images. Mean and standard deviation of ΔS over all patients are reported. A negative value indicates that the acquired signal intensity is higher than the synthesized signal intensity. Further, violin plots were generated for the 50^th^, 95^th^, and 98^th^ percentile of signal intensity within several ROIs: prostate; prostate plus margin (5 mm, 30 mm, or 70 mm); and the whole FOV. For the whole FOV, values higher than 3000 signal intensity units (SIU) were capped and set to 3000 SIU. Violin plots present the median value in combination with the kernel density distribution ^31^.

Lesion conspicuity was evaluated using the contrast-to-noise ratio (CNR) between lesion and surrounding prostate tissue. CNR is defined as the following:

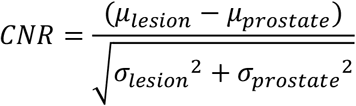

where μ is the mean signal of the ROI under consideration and σ the standard deviation. CNR was evaluated for all patients and patients diagnosed with csPCa. A higher CNR indicates a better tumor conspicuity^32^. Significant differences between CNRs of different images and patient cohorts were tested using two sample t-test with a confidence level of 0.01.

Prediction of whether csPCa was found on biopsy was also evaluated for aDWI, sDWI, and RSIrs. RSIrs is a quantitative cancer biomarker based on a multi-exponential DWI model and has been previously shown to be more accurate than conventional DWI^20–22^. Computation of RSIrs for this dataset was performed previously and is described in detail in previous publications^21,22,33^. Briefly, the coefficient for the slowest diffusion compartment (corresponding to intracellular restricted diffusion) was normalized by the median signal within the prostate on *b*=0 s/mm^2^ images. The maximum aDWI, sDWI, or RSIrs value within each considered ROI was used as the predictor variable, as done in previous work^22^. This is analogous to the maximum standard uptake value (SUV) in quantitative Positron Emission Tomography (PET) imaging. Receiver-operating characteristic (ROC) curves were calculated, and the area under the curve (AUC) reported for aDWI, sDWI, and RSIrs. The false positive rate at 90% sensitivity (FPR90) was also reported for each metric. AUC and FPR90 were compared using bootstrap (N=10,000) 95% confidence intervals and *p*-values.

## Results

**Figure 1** shows the difference between acquired and synthesized *b*-values for a representative patient using the mean signal intensity within the prostate. Within the prostate, mean ± standard deviation of percent differences between sDWI and aDWI were -46±35% for sDWI_1000_ and -67±24% for sDWI_500_. A negative error indicates sDWI had lower values than aDWI. Comparing sDWI_1000_ to sDWI_500_, a difference of -41±4% was estimated (see **Table 3**). sDWI_500_ had overall larger errors than sDWI_1000_. Signal intensity of aDWI was lower than sDWI in the prostate and in the prostate plus 5 mm margin, as indicated by a negative mean difference and lower median for the 50^th^, 95^th^ and 98^th^ percentiles. The 50^th^ percentile of aDWI is higher than sDWI for all considered ROIs. For the 95^th^ and 98^th^ percentiles, however, sDWI is larger for margins of ≥30 mm beyond the prostate. The standard deviation of sDWI is larger than that of aDWI for all ROIs and all considered percentiles. Comparison of the 50^th^, 95^th^ and 98^th^ percentiles for five ROIs is shown in **Figure 3**. Mean and standard deviation of CNR over all patients was 0.95±0.87, 0.84±0.80, 0.65±0.66 and 0.97±0.79 for aDWI, sDWI_1000,_ sDWI_500_ and RSIrs, respectively. A lower CNR indicates a lower tumor conspicuity. CNR considering only patients with csPCa changed to 1.00±0.84, 0.86±0.78, 0.65±0.65 and 0.99±0.76 for aDWI, sDWI_1000,_ sDWI_500_ and RSIrs respectively. CNR for aDWI and RSIrs proved to be significantly different to sDWI (p<0.01) for all patients and patients with csPCa. **Figure 5** compares sDWI, aDWI and RSIrs for detection of significant cancer lesions outside of the prostate.

**Table 3.** Summary of median differences between synthesized (sDWI) and acquired (aDWI) diffusion-weighted imaging. Comparison between sDWI and aDWI is presented relative to aDW. FOV= field of view

| ROI | Datasets | Median Difference ± Interquartile range | | |
| --- | --- | --- | --- | --- |
|  |  | Total (N = 151) | csPCA (N = 86) | Benign (N = 65) |
| Prostate | sDWI<sub>1000</sub> vs. aDWI | -55±27% | -52±29% | -59±22% |
| | sDWI<sub>500</sub> vs. aDWI | -72±15% | -69±18% | -74±13% |
| Prostate+5 mm margin | sDWI<sub>1000</sub> vs. aDWI | -55±26% | -53±29% | -58±21% |
| | sDWI<sub>500</sub> vs. aDWI | -72±18% | -69±19% | -74±13% |
| Prostate+30 mm margin | sDWI<sub>1000</sub> vs. aDWI | -48±35% | -46±41% | -50±28% |
| | sDWI<sub>500</sub> vs. aDWI | -58±34% | -54±45% | -61±23% |
| Prostate+70 mm margin | sDWI<sub>1000</sub> vs. aDWI | 1.9e19±4.1e19% | 1.8e19±3.8e19% | 2.0e19±6.4e19% |
| | sDWI<sub>500</sub> vs. aDWI | 1.5e47±1.7e48% | 1.4e47±1.2e48% | 1.5e47±3.0e48% |
| Whole FOV | sDWI<sub>1000</sub> vs. aDWI | 1.1e19±2.3e19% | 1.1e19±2.2e19% | 1.6e19±2.5e19% |
| | sDWI<sub>500</sub> vs. aDWI | 3.4e48±8.9e50% | 2.0e48±7.1e50% | 1.3e49±9.2e50% |

**Figure 1.**
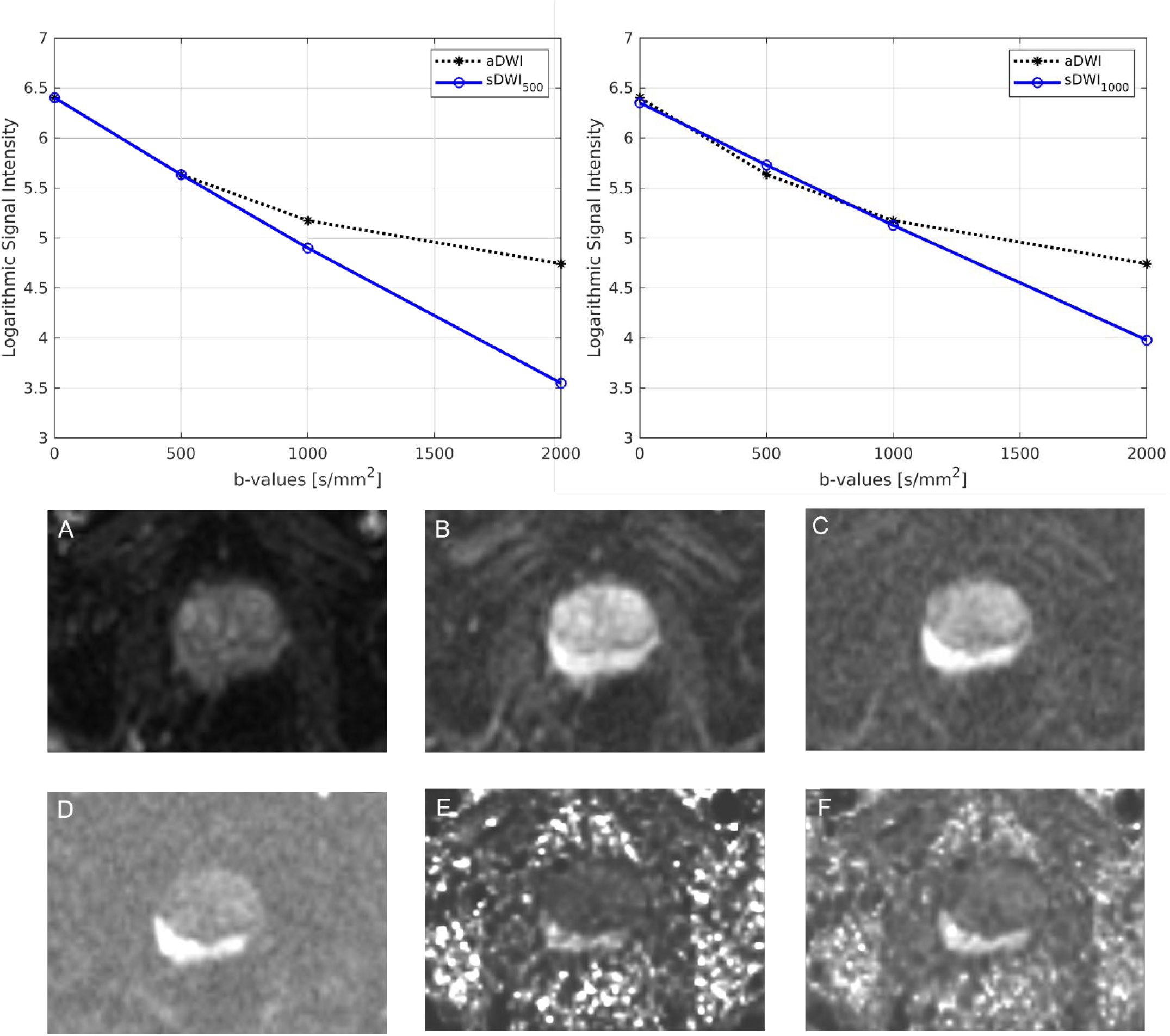
Comparison of acquired images to those synthesized with mono-exponential models using either b-values up to 500 s/mm^2^ (sDWI_500_) or b-values up to 1000 s/mm^2^ (sDWI_1000_) presented for one representative patient. Mean value within the prostate are compared. sDWI is not an accurate representation of aDWI at b=2000. Figures A-F show the different diffusion images for one patient. A-D present the acquired images for b=0 s/mm^2^ (A), b=500 s/mm^2^ (B), b=1000 s/mm^2^ (C) and b=2000 s/mm^2^ (D). E and F show the synthesized b=2000 s/mm^2^ images. E shows sDWI_500_ and F sDWI_1000_. aDWI = acquired diffusion-weighted image for b=2000 s/mm^2^

**Figure 2.**
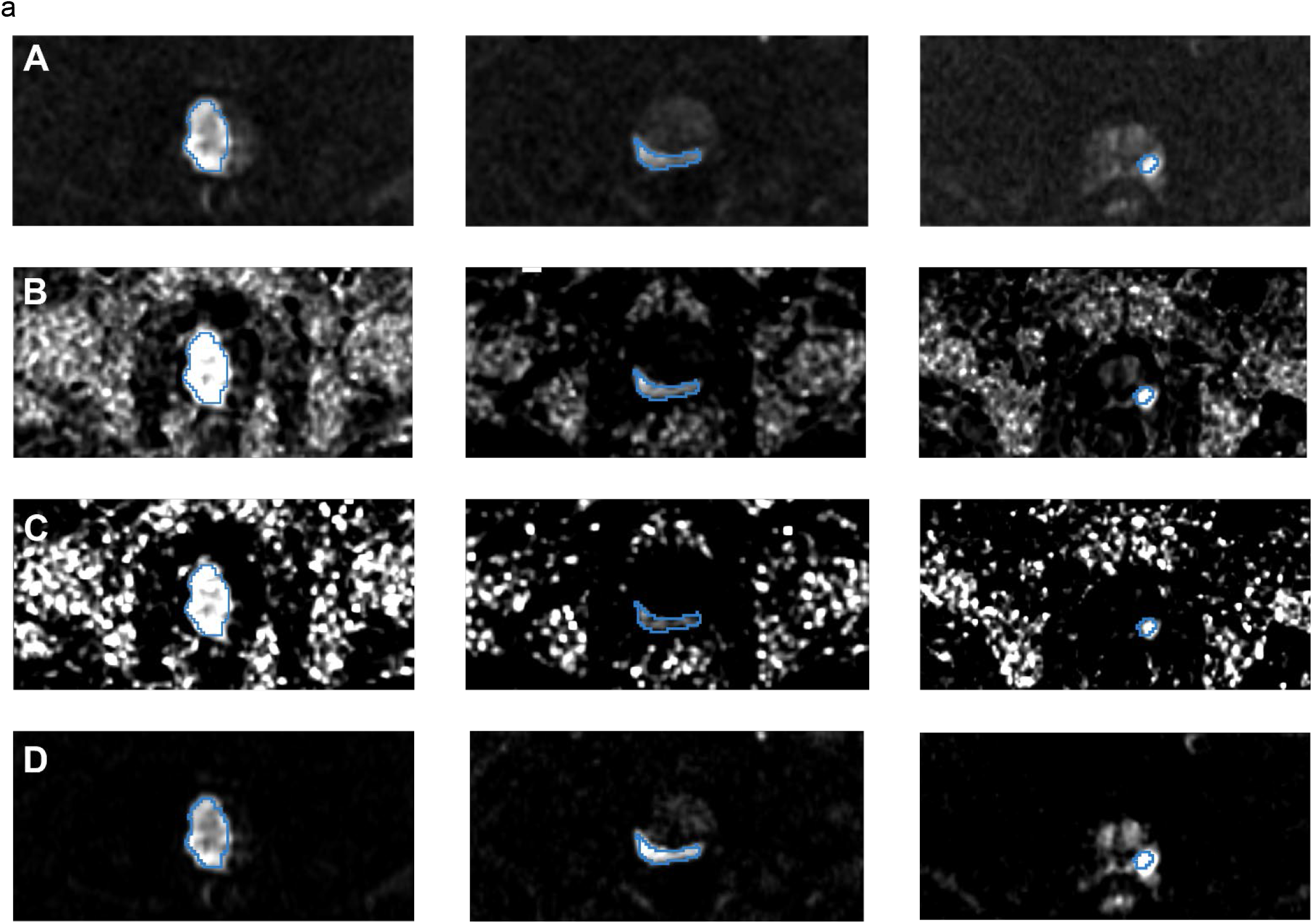
Representative images from three patients (corresponding to three columns). (A) acquired diffusion-weighted image (aDWI) for b=2000 s/mm^2^, (B) synthesized DWI using acquired b-values up to b=1000 s/mm^2^ (sDWI_1000_), (C) synthesized DWI using acquired values up to b=500 s/mm^2^ (sDWI_500_), and (D) restriction spectrum imaging restriction score (RSIrs). The radiologist-defined cancer lesion for each patient is indicated in blue. All presented patients had a PI-RADS score of 5. The same window level was chosen for all presented images.

For detection of csPCa, the AUCs for sDWI and aDWI were similar in both the prostate and prostate plus 5 mm (**Figure 3** and **Table 4**). Classification accuracy decreased significantly for sDWI when considering the whole FOV (AUC = 0.45 [0.36, 0.54] for sDWI_1000_ and 0.47 [0.38, 0.56] for sDWI_500_). RSIrs was superior to sDWI and aDWI for all ROIs (*p*<0.01). The AUC of RSIrs was 0.77 [0.69, 0.84] within prostate plus 5 mm and decreased to 0.70 [0.61, 0.78] for the whole FOV. FPR90 was similar for aDWI and sDWI in all ROIs. Mean FPR90 was significantly lower for RSIrs than for either aDWI or sDWI, indicating fewer false positives (*p*<0.05).

**Table 4.** Classification accuracy for the detection of cancer is shown for the prostate, prostate plus a 5 mm margin and the whole field of view (FOV). For statistical comparison bootstrapping (N=10,000) was performed and the 95% confidence intervals (CI) of AUC and the mean false positive rate at 90% sensitivity (FPR90) reported. ROI = Region of interest; AUC = area under the curve; RSIrs = biomarker based on restriction spectrum imaging; aDWI = acquired diffusion b=2000 s/mm^2^ MRI; sDWI_1000_ = synthesized image using acquired b-values up to 1000s/mm^2^; sDWI_500_ = synthesized image using acquired b-values up to 500 s/mm^2^; * significantly different with p < 0.05 in comparison to each of the other metrics

| ROI | Dataset | AUC | 95<sup>th</sup> CI | FPR90 |
| --- | --- | --- | --- | --- |
| Prostate | RSIRS | 0.78* | 0.71-0.86 | 0.50* |
|  | aDWI | 0.62 | 0.53-0.71 | 0.82 |
|  | sDWI<sub>1000</sub> | 0.65 | 0.56-0.73 | 0.76 |
|  | sDWI<sub>500</sub> | 0.63 | 0.54-0.72 | 0.76 |
| Prostate +<br>5 mm<br>margin | RSIRS | 0.77* | 0.68-0.84 | 0.53* |
|  | aDWI | 0.61 | 0.51-0.69 | 0.87 |
|  | sDWI<sub>1000</sub> | 0.60 | 0.51-0.69 | 0.84 |
|  | sDWI<sub>500</sub> | 0.56 | 0.47-0.65 | 0.84 |
| Prostate +<br>30 mm<br>margin | RSIRS | 0.72* | 0.64-0.80 | 0.62* |
|  | aDWI | 0.58 | 0.49-0.67 | 0.89 |
|  | sDWI<sub>1000</sub> | 0.52 | 0.43-0.61 | 0.83 |
|  | sDWI<sub>500</sub> | 0.51 | 0.42-0.61 | 0.86 |
| Prostate +<br>70 mm<br>margin | RSIRS | 0.72* | 0.64-0.81 | 0.61* |
|  | aDWI | 0.60 | 0.50-0.69 | 0.88 |
|  | sDWI<sub>1000</sub> | 0.46 | 0.36-0.55 | 0.87 |
|  | sDWI<sub>500</sub> | 0.45 | 0.36-0.55 | 0.89 |
| Whole FOV | RSIRS | 0.70* | 0.61-0.78 | 0.62* |
|  | aDWI | 0.59* | 0.49-0.68 | 0.90 |
|  | sDWI<sub>1000</sub> | 0.45 | 0.36-0.54 | 0.91 |
|  | sDWI<sub>500</sub> | 0.47 | 0.38-0.56 | 0.90 |

**Figure 3.**
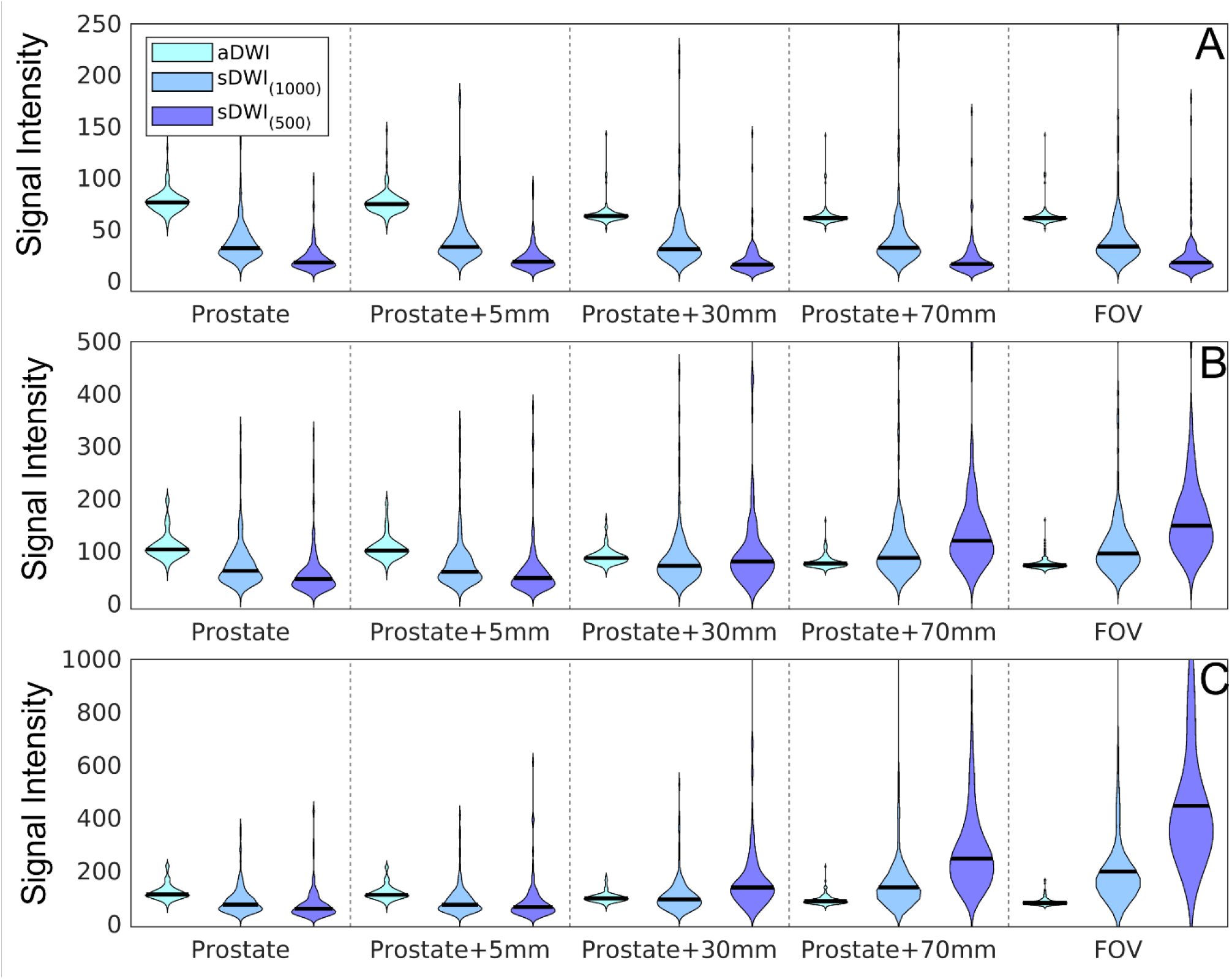
Violin plots summarizing the signal intensity across 151 patients for (A) 50^th^ percentile, (B) 95^th^ percentile, (C) and 98^th^ percentiles of various DWI metrics calculated for each patient. The percentiles are estimated over different regions of interest: the prostate; the prostate with varying margin (5 mm, 30 mm, or 70 mm); and the whole field of view. aDWI = acquired diffusion-weighted image with b=2000 s/mm^2^; sDWI = synthesized DWI for b=2000 s/mm^2^ using either acquired b-values up to 1000 s/mm^2^ (sDWI_1000_) or up to 500 s/mm^2^ (sDWI_500_).

**Figure 4.**
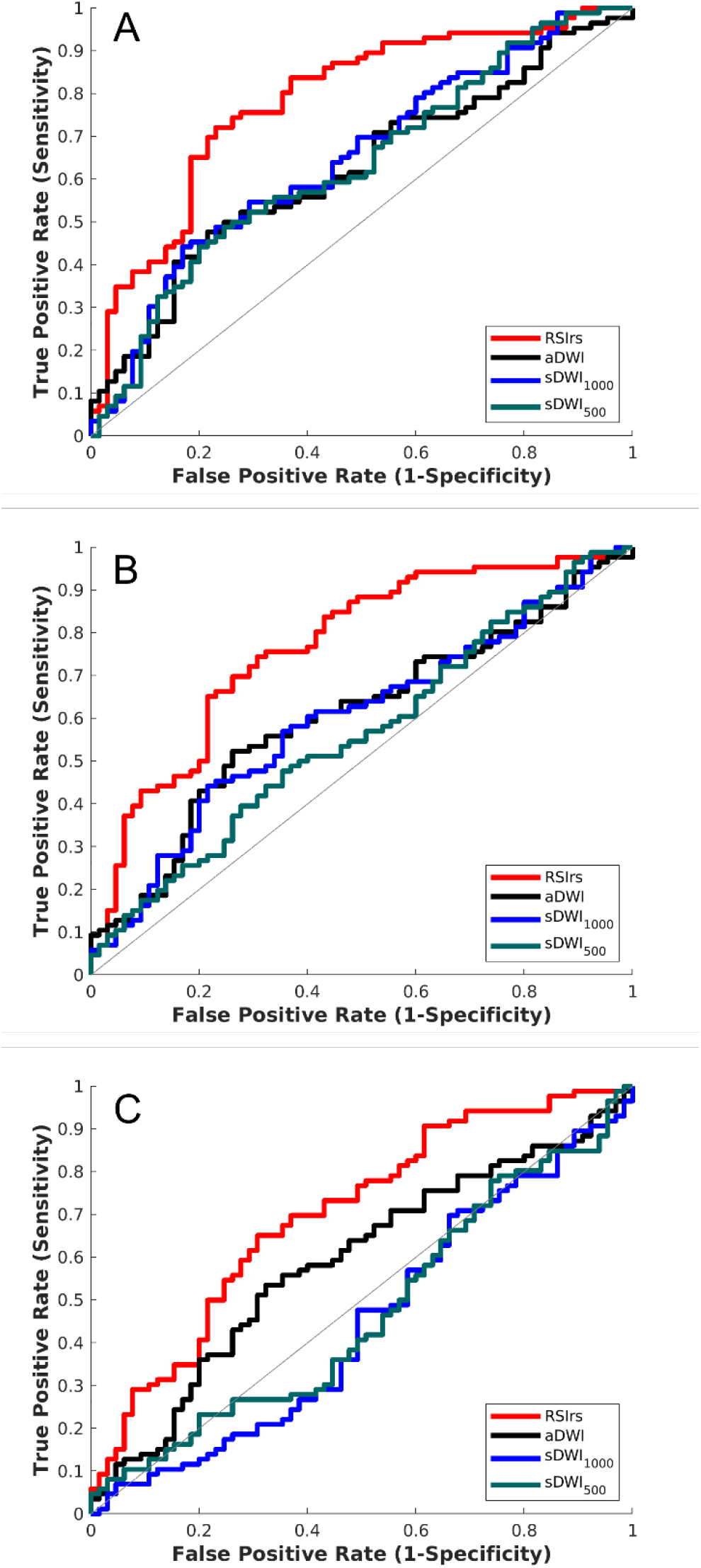
ROC curves for DWI metrics within three ROIs (A) the prostate, (B) the prostate with 5 mm margin, and (C) the whole field of view. The DWI metrics compared for classification accuracy are RSIrs, acquired diffusion-weighted images (aDWI), synthesized DWI using b-values up to 1000 s/mm^2^ (sDWI_1000_), and synthesized DWI using b-values up to 500 s/mm^2^ (sDWI_500_). Clinically significant prostate cancer was defined as a Gleason score ≥2.

## Discussion

We found that synthesized DWI images can be qualitatively similar to acquired DWI within the prostate even though sDWI is quantitatively an inaccurate representation of aDWI. Moreover, sDWI introduces unacceptable artifacts and inaccuracies in pelvic tissues. sDWI_500_ calculated using only *b*-values up to 500 s/mm^2^ was inferior to sDWI_1000_. Acquisitions of an increased number of lower *b*-values might have the possibility to improve sDWI but would potentially increase the scan time. sDWI proved to be systematically different from aDWI within the prostate and in the whole FOV. Even within the prostate, sDWI and aDWI differed between 55-72%. Nonetheless, within the prostate and the prostate plus 5 mm margin, lesion conspicuity was reasonably preserved. Both sDWI and aDWI had a similar quantitative performance in detecting csPCa with an AUC ranging between 0.56-0.65. However, sDWI introduced larger errors in the surrounding pelvic tissue even in a reduced FOV acquisition. Overall RSIrs outperformed sDWI and aDWI for quantitative prediction of biopsy-proven csPCa for all considered ROIs.

Severe artifacts were observed on sDWI in the pelvic tissue outside of the prostate, in particular for sDWI_500_, which makes the detection of metastasis outside of the prostate region difficult, **Figure 5**. There are many ways to potentially improve the calculation of synthesized images including bi-exponential or multi-exponential modeling^23^. For example, RSIrs is based on a multi-exponential model and may synthesize images without introducing artifacts. Image artifacts may be explained by poor signal quality, magnitude smaller than one in a subset of voxels, or noise/distortion correction post image acquisition leading to voxels with very low signal intensity. In particular, mono-exponential models fail to correctly represent voxels with low signal intensity due to exponential fitting. Smoother images could be created by censoring those voxels by interpolating from surrounding voxels, smoothing low *b-*value images prior to calculation, or by thresholding low intensity voxels. For quantitative imaging, the details of such decisions would need to be clearly described and accounted for, and potentially could lead to more false positive/negative detections. Such enhanced images would not represent the measured truth and would include some unreliable voxels, which must be taken into consideration when interpreting the images. A “nicer looking” image does not necessarily mean that the image quality or reliability is better.

**Figure 5.**
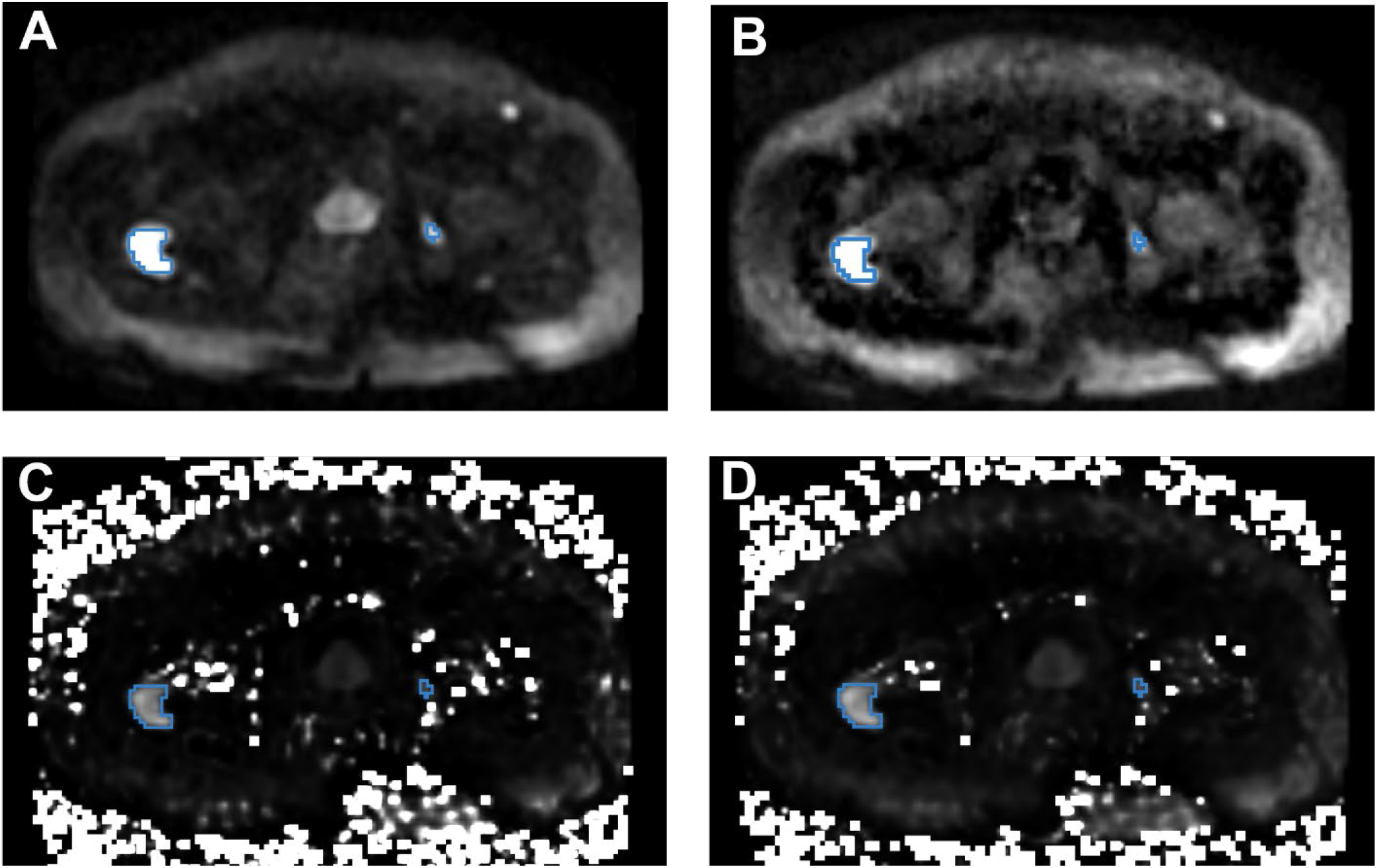
An example of pelvic DWI in a patient with prostate cancer bone metastases. The blue contours mark the cancer lesions. (A) acquired DWI with b=2000 s/mm2; (B) RSIrs based on restriction spectrum imaging; (C) synthesized DWI using acquired b-values up to 500 s/mm^2^; and (D) synthesized DWI using acquired b-values up to 1000 s/mm^2^. Tumor lesions are easily detectible in A and B, but there is more high-intensity artifact on sDWI, and the smaller bone metastasis is not as easily identifiable in C and D.

Prior studies reported sDWI to have higher subjective quality and tumor conspicuity^5,27^. This may reflect the particular imaging sequences and, platforms used, or the particular image enhancement effects. It is also important to note that these prior results were mostly subjective judgements and not quantitative assessments of accuracy. In the presented study we have proven that tumor conspicuity is quantitatively greater with aDWI (CNR = 0.95) in comparison to sDWI (CNR = 0.65-0.84). Another option to improve sDWI is to use a multi-compartment DWI model, e.g., RSIrs, for a more stable and accurate signal extrapolation. For example, RSIrs outperformed both sDWI and aDWI in the present study. Other promising multi-compartment models have proved to be superior to conventional multiparametric MRI, like a biomarker derived from VERDICT that outperformed ADC in the detection of csPCa^14^. Hybrid multidimensional MRI acquisitions also showed promising results for classifying csPCa with a reported AUC of 0.94^34^.

One limiting factor of our study was that we only considered a retrospective dataset form a single scanner and a single institution. Further, only the conventional mono-exponential model was tested in the presented study. A precise comparison of all possible methods for synthesizing DWI is beyond the scope of this manuscript. RSIrs is one quantitative biomarker based on a multi-compartment model. The acquisition protocol in the datasets here was not optimized for models like hybrid multi-dimensional MRI or VERDICT.

### Conclusion

Within the prostate, sDWI is a systematically inaccurate representation of aDWI, but the techniques are quantitatively comparable in terms of detecting csPCa. In the surrounding pelvic tissue, high signal intensity artifacts are introduced with sDWI. These artifacts decrease the csPCa detection rate in surrounding tissues and might mask potential metastases within the pelvis. RSIrs is superior to either sDWI or aDWI for quantitative csPCa detection. Despite the quantitative inaccuracies, sDWI may still be adequate for current subject clinical interpretation within the prostate.

## Data Availability

All data produced in the present study are available upon reasonable request to the authors

## Abbreviation

DWI: diffusion-weighted imaging
csPCa: clinically significant prostate cancer
FOV: field of view
RSIrs: restriction spectrum imaging restriction score
CNR: contrast-to-noise ratio
sDWI: synthesized diffusion-weighted image
aDWI: aquired diffusion-weighted image
ROI: region of interest
ROC: receiver operating characteristic
AUC: area under the curve
FPR90: false positive rate at 90% sensitivity
SNR: signal to noise ratio

